# Resveratrol and Copper for treatment of severe COVID-19: an observational study (RESCU 002)

**DOI:** 10.1101/2020.07.21.20151423

**Authors:** Indraneel Mittra, Rosemarie de Souza, Rakesh Bhadade, Tushar Madke, P.D. Shankpal, Mohan Joshi, Burhanuddin Qayyumi, Atanu Bhattacharjee, Vikram Gota, Sudeep Gupta, Pankaj Chaturvedi, Rajendra Badwe

**Affiliations:** Tata Memorial Centre, Mumbai, India; Homi Bhabha National Institute, Mumbai, India; BYL Nair Charitable Hospital, Mumbai, India

## Abstract

**Background:** To be universally applicable in treatment of severe COVID-19, novel therapies, especially those with little toxicity and low cost, are urgently needed. We report here the use of one such therapeutic combination involving two commonly used nutraceuticals, namely resveratrol and copper in patients with this disease. This study was prompted by pre-clinical reports that sepsis-related cytokine storm and fatality in mice can be prevented by oral administration of small quantities of resveratrol and copper. Since cytokine storm and sepsis are major causes of death in severe COVID-19, we retrospectively analyzed outcomes of patients with this condition who had received resveratrol and copper.

**Methods & Findings:** Our analysis comprised of 230 patients with severe COVID-19 requiring inhaled oxygen who were admitted in a single tertiary care hospital in Mumbai between April 1 and May 13 2020. Thirty of these patients received, in addition to standard care, resveratrol and copper at doses of 5.6 mg and 560 ng, respectively, orally, once every 6 hours, until discharge or death. These doses were based on our pre-clinical studies, and were nearly 50 times and 2000 times less, respectively, than those recommended as health supplements. A multivariable-adjusted analysis was used to model the outcome of death in these patients and evaluate factors associated with this event. A binary logistic regression analysis was used, with age, sex, presence of comorbidities and receipt of resveratrol-copper as covariates. Data were updated as of May 30 2020. The number of deaths in resveratrol-copper and standard care only groups were 7/30 (23.3%, 95% CI 8.1%-38.4%) and 89/200 (44.5%, 95% CI 37.6%-51.3%), respectively. In multivariable analysis, age >50 years [odds ratio (OR) 2.558, 95% CI 1.454-4.302, *P*=0.0011] and female sex (OR 1.939, 95% CI 1.079-3.482, *P*=0.0267) were significantly associated, while presence of co-morbidities was not significantly associated (OR 0.713, 95% CI 0.405-1.256, *P*=0.2421) with death. There was a trend towards reduction in death in patients receiving resveratrol-copper (OR 0.413, 95% CI 0.164-1.039, *P*= 0.0604).

**Conclusions:** We provide preliminary results of a novel approach to the treatment of severe COVID-19 using a combination of small amounts of commonly used nutraceuticals, which is non-toxic and inexpensive, and therefore could be widely accessible globally. The nearly two-fold reduction in mortality with resveratrol-copper observed in our study needs to be confirmed in a randomized controlled trial.

## Introduction

It has been previously reported that cell-free chromatin particles that are released from the billions of cells that die in the body everyday can illegitimately integrate into genomes of healthy cells to trigger DNA double-strand breaks, apoptosis and release of inflammatory cytokines^**1-6**^. Based on these findings we hypothesized that cell-free chromatin particles that are released from dying host cells following SARS-COV-2 infection could integrate into genomes of uninfected cells to trigger more host cell death leading to a vicious cycle of further rounds of DNA damage, apoptosis and inflammation. These events could perpetuate and amplify the pathological effects of SARS-COV-2 infection. This hypothesis was supported by our preclinical study which demonstrated that endotoxin induced cytokine storm and sepsis related fatality in mice can be prevented by oral administration of small quantities of resveratrol and copper^**7**^. Since cytokine storm and sepsis are major causes of death in severe COVID-19^**8**^, we analyzed the efficacy of R-Cu in preventing death from this condition.

Resveratrol is an anti-oxidant nutraceutical plant polyphenol which has been extensively researched for its health benefits^**9**^. It has been reported that resveratrol acts as a pro-oxidant in presence of copper ^**10**^ which is another widely researched nutraceutical ^**11**^. Resveratrol can reduce copper (II) to copper (I) thereby generating highly unstable free-radicals ^**10**^ which can degrade cell-free chromatin ^**7, 12-14**^ and can lead to prevention of endotoxin sepsis in mice ^**7**^. The pro-oxidant activity of resveratrol-copper is maintained even when the molar concentration of copper is reduced 10,000 fold ^**7, 12-14**^. Doses of resveratrol and copper taken by the patients included in this analysis were nearly 50 times and 2000 times less, respectively, than those currently recommended as health supplements ^**15, 16**^.

## Methods

### Patient Characteristics

Patients with severe COVID-19 requiring oxygen therapy and who had been admitted to B. Y. L. Nair Charitable Hospital and TN Medical College, Mumbai, under a single faculty member (RD) were provided access to resveratrol-copper. All patients had dyspnea, oxygen saturation of ≤ 92% at room air and lung opacities on chest radiographs. Patient characteristics are described in Table 1.

**Table 1:** Patient Characteristics and Deaths Related to Receipt of Resveratrol-copper

| Variables | Entire Cohort<br>(N = 230)<br>n (%) | Resveratrol -<br>copper<br>No<br>(N = 200)<br>n (%) | Resveratrol<br>– copper<br>Yes<br>(N = 30)<br>n (%) | P-Value |
| --- | --- | --- | --- | --- |
| **Age** |  |  |  |  |
| Mean ± SD | 51 ±15.40 | 51.3 ±15.96 | 49.16 ± 11 | 0.2752 |
| Median | 52 | 52.5 | 48.5 |  |
| ≤ 50 | 109 (47.4%) | 92 (46%) | 17 (56.7%) |  |
| > 50 | 121 (52.6%) | 108 (56%) | 13 (43.3%) |  |
| **Sex** |  |  |  |  |
| Male | 155 (67.4%) | 135 (67.5%) | 20 (66.7%) | 0.9277 |
| Female | 75 (32.6%) | 65 (32.5%) | 10 (33.3%) |  |
| **Comorbidities\*** |  |  |  |  |
| No | 109 (47.4%) | 99 (49.5%) | 10 (33.3%) | 0.0982 |
| Yes | 121 (52.6%) | 101 (50.5%) | 20 (66.7%) |  |
| **Death** |  |  |  |  |
| No | 134 (58.3%) | 111 (55.5%) | 23 (76.7%) | 0.0284 |
| Yes | 96 (41.7%) | 89 (44.5%) | 7 (23.3%) |  |

**\*Comorbidities:** Diabetes, hypertension, malignancy, hypothyroidism, chronic kidney disease, cardio-vascular accident, Ischemic heart disease, bronchitis, retroviral disease, asthma, chronic obstructive pulmonary disease, Koch's (under-treatment), dengue, atrial fibrillation, RH incompatibility and anaemia.

### Standard of Care

All patients received standard of care as follows: high flow oxygen which was administered through nasal prongs or face masks with properly fitted circuits or through high flow oxygen nasal cannulas. The flow rates were adjusted to deliver adequate oxygenation, and awake proning was performed. If patients deteriorated, they were intubated and put on a ventilator. Patients were administered oral hydroxychloroquine 400 mg twice a day on day 1 followed by 400 mg once a day for 4 days. Azithromycin 500 mg was administered once a day for 5 days, and vitamin D 60,000 units once per week for 6 weeks and vitamin C 500 mg once a day. Low molecular weight heparin or unfractionated heparin in therapeutic doses was given to all patients with serum D–dimer levels of > 6, while prophylactic anticoagulation with either low molecular weight heparin or unfractionated heparin was administered to patients with D-dimer levels between 3 and 6. Patients with oxygen requirement of >6 litres per minute were also administered injectable methylprednisolone at a dose of 40mg twice a day for short durations. Treatment of comorbidities like diabetes and hypertension were treated appropriately. Diabetes was treated with insulin.

### Resveratrol-Copper Intervention

Trans-resveratrol was procured from Biotivia LLC, USA, and chelated copper from J R Carlson Laboratories Inc., USA. Resveratrol (5.6 mg) was suspended in 20 mL of water and administered orally followed by 20 mL of water as wash down. Copper (560 ng) in 20 mL of water-based solution was administered orally immediately afterwards followed by 20 mL of water as wash down. This resveratrol-copper treatment was given every 6 hours until the patient was discharged or died. These human doses of resveratrol and copper were arrived at by direct conversion of doses employed in our pre-clinical studies using a conventional formula ^1**7**^.

### Study Design & Outcome Measures

Clinical outcome of patients receiving resveratrol-copper was compared with 200 contemporaneously treated patients with severe COVID-19 and same clinical criteria, at the same institution. The primary outcome measure was death due to any cause within 28 days of diagnosis.

### Statistical Analysis

The primary analysis was a binary logistic regression with inclusion of age, sex, presence of comorbidities (as a single variable), and use of resveratrol-copper as covariates in all patients fulfilling eligibility criteria. The likelihood ratio was utilized to examine the significance of covariates included in the model with a two-sided *P*-value of less than 0.05 to indicate statistical significance. Additional analyses were performed to compare the proportion of deaths within subgroups defined by age, sex, presence of comorbidities and use of resveratrol-copper, using chi-square test. All statistical analyses were performed with R 3.6.3 software (https://cran.r-project.org/).

### Study Oversight

This observational study was approved by the Ethics Committee of BYL Nair Hospital, Mumbai. The study is registered in the Clinical Trials Registry of India; Registration Number CTRI/2020/06/026256 The analysis was conducted adhering to ethical principles, including maintaining patient confidentiality. All authors vouch for the accuracy of data and analysis.

## Results

### Participants

Between April 1 and May 13, 2020, 241 patients fulfilling the eligibility criteria for this analysis were admitted, of whom 32, admitted under a single faculty member, received resveratrol-copper (Table 1). Of these 241 patients, nine receiving standard treatment and one patient receiving resveratrol-copper took discharge from the hospital against medical advice, while one patient receiving resveratrol-copper was transferred to another hospital for renal dialysis. The current analysis includes remaining 230 patients of whom 30 patients received resveratrol-copper.

### Demographics

Mean ages (±SD) of the patients were 49.16 ±11 years and 51.3 ±15.96 years in the resveratrol-copper and control groups, respectively; while corresponding proportions of female patients were 10 (33.3%) and 65 (32.5%), and patients with comorbidities were 20 (66.7%) and 101 (50.5%), respectively. The complete set of de-identified data used in this analysis will be provided on request.

### Outcome

At the time of analysis, 7 of 30 patients (23.3%) who had been treated with resveratrol-copper and 89 of 200 patients (44.5%) who had not received resveratrol-copper had died [odds ratio (OR) of death, 0.38; 95% confidence interval (CI), 0.156 to 0.925; *P*=0.0284] (<u>Table</u> 1). In multivariable analysis, increasing age was the most significant adverse prognostic factor (OR of death, 2.558; 95% CI, 1.456 to 4.495; *P*=0.001) (<u>Table 2</u>). Female sex was also a significant adverse factor (OR of death, 1.939; 95% CI, 1.079 to 3.482; *P*=0.026). Death due to co-morbidities failed to reach statistical significance (OR of death, 0.713, 95% CI 0.405-1.256, *P*=0.2421). Treatment with resveratrol-copper showed a trend towards reducing deaths (OR of death, 0.413; 95% CI, 0.164 to 1.039; *P*=0.0604) (Table 2).

**Table 2:** Univariate and Multivariable Analyses of Factors Impacting Death

|  | **Death n (%)** | **Univariate Odds Ratio (95% CI)** | **Univariate *P*-value** | **Multivariable Odds Ratio (95% CI)** | **Multi-variable *P*-value** |
| --- | --- | --- | --- | --- | --- |
| **Age** |  |  |  |  |  |
| ≤ 50 | 33/109 (30.3%) |  |  |  |  |
| vs |  | 2.501 | 0.0008 | 2.558 | 0.0011 |
| >50 | 63/121 (52.1%) | (1.454, 4.302) |  | (1.456, 4.495) |  |
| **Sex** |  |  |  |  |  |
| Male | 57/155 (36.8%) |  |  |  |  |
| vs |  | 1.863 | 0.0282 | 1.939 | 0.0267 |
| Female | 39/75 (52.0%) | (1.066, 3.255) |  | (1.079, 3.482) |  |
| **Comorbidities** |  |  |  |  |  |
| No | 48/109 (44%) |  |  |  |  |
| vs |  | 0.836 | 0.5024 | 0.713 | 0.2421 |
| Yes | 48/121 (39.7%) | (0.494, 1.413) |  | (0.405, 1.256) |  |
| **Resveratrol - copper Given** |  |  |  |  |  |
| No | 89/200 (44.5%) |  |  |  |  |
| vs |  | 0.38 | 0.0284 | 0.413 | 0.0604 |
| Yes | 7/30. (23.3%) | (0.156, 0.925) |  | (0.164, 1.039) |  |

## Discussion

The results of this retrospective study suggest that treatment with a combination of the nutraceuticals resveratrol and copper administered in minuscule quantities could lead to a nearly 2-fold reduction in mortality in patients suffering from severe COVID-19 and associated ARDS. The mechanism of action of resveratrol-copper in COVID-19 patients is unclear, but could be related to the generation of free radicals which can inactivate or degrade cell-free chromatin released from dying cells and contributing to the sepsis cascade. Not being a randomized clinical trial, results of this study should be considered as hypothesis generating rather than being confirmatory. Nonetheless, to our knowledge this is one of the few studies to show that a therapeutic intervention can prevent death from this serious condition.

The overall mortality in admitted patients in our series is 41.7%, which is higher than that reported in other series^**18**,**19**^. This is likely because many patients arrived at the hospital in severe sepsis and advanced hypoxemia and died rapidly after admission. We also observe, like almost all other series, that increasing age and presence of comorbidity increases the odds of death. However, unlike others, our analysis suggests that female patients have a higher risk of dying compared with men, even after adjusting for age and presence of comorbidity. This has also been noted in other analyses from India ^**19**^ but the reasons are unclear. It is possible that, because of socioeconomic reasons, female patients are brought to the hospital when they have very severe disease and a disease-severity adjusted analysis will overcome this confounding. Nutritional deficiencies like iron deficiency and others are also more prevalent in Indian women and could play a role in this observation.

In conclusion, our results of treating severe COVID-19 patients with a combination of the nutraceuticals resveratrol and copper are promising which need to be confirmed in a randomized clinical trial.

## Data Availability

All de-identified patient data used for analysis in this study is made available as table 2 in the manuscript.

## References

1. Raghuram GV, Chaudhary S, Johari S et al. Illegitimate and repeated genomic integration of cell-free chromatin in the aetiology of somatic mosaicism, ageing, chronic diseases and cancer. Genes 2019; 10: 407–426.

2. Mittra I., Khare NK, Raghuram GV et al. Circulating nucleic acids damage DNA of healthy cells by integrating into their genomes. Journal of Biosceince 2018; 40: 91–111.

3. Basak R, Nair NK & Mittra I. Evidence for cell-free nucleic acids as continuously arising endogenous DNA mutagens. Mutat. Res. - Fundam. Mol. Mech. Mutagen 2016; 794: 15–21.

4. Mittra I, Samant U, Sharma S et al. Cell-free chromatin from dying cancer cells integrate into genomes of bystander healthy cells to induce DNA damage and inflammation. Cell Death Discov 2017; 3: 1–14.

5. Chaudhary S, Raghuram, G. & Mittra I. Is inflammation a direct response to dsDNA breaks? Mutat. Res. - Fundam. Mol. Mech. Mutagen 2018; 808: 48–52.

6. Chaudhary S. & Mittra I. Cell-free chromatin: A newly described mediator of systemic inflammation. J. Bioscience 2019; 44: 32–38.

7. Mittra I, Pal K, Pancholi N et al. Cell-free chromatin particles released from dying host cells are global instigators of endotoxin sepsis in mice. PLoS One 2020; 15: 1–22.

8. Bellinvia S, Edwards CJ, Schisano M, et al. The unleashing of the immune system in COVID-19 and sepsis: the calm before the storm? Inflamm. Research 2020; 1-7.

9. Diaz-Gerevini GT, Repossi G, Dain A, et al. Beneficial action of resveratrol: How and why? Nutrition 2016; 32: 174–178.

10. Fukuhara K & Miyata N. Resveratrol as a new type of DNA-cleaving agent. Bioorganic Med. Chem. Lett 1998; 8, 3187–3192.

11. Bost M, Houdart S, Oberli M, et al. Dietary copper and human health: Current evidence and unresolved issues. Journal of Trace Elements in Medicine and Biology 2016; 35: 107–115.

12. Subramaniam S, Vohra I, Iyer A, et al. A paradoxical relationship between resveratrol and copper (II) with respect to degradation of DNA and RNA. F1000 Research 2015; 4: 1145–1158.

13. Mittra I, Pal K, Pancholi N et al. Prevention of chemotherapy toxicity by agents that neutralize or degrade cell-free chromatin. Ann. Oncol 2017; 28: 2119– 2127.

14. Kirolikar S, Prasannan P, Raghuram GV et al. Prevention of radiation-induced bystander effects by agents that inactivate cell-free chromatin released from irradiated dying cells. Cell Death Disease 2018; 9:1142–1158.

15. TRANSMAX™ | Biotivia. https://www.biotivia.com/product/transmax/

16. Chelated Copper https://carlsonlabs.com/chelated-copper/

17. Reagan-Shaw S, Nihal M and Ahmad N. Dose translation from animal to human studies revisited. FASEB J 2008; 22: 659–661.

18. Grein J, Ohmagari N, Shin D, et al. Compassionate Use of Remdesivir for Patients with Severe Covid-19. N Engl J Med 2020;382:2327–36.

19. Goldman JD, Lye DCB, Hui DS, et al. Remdesivir for 5 or 10 Days in Patients with Severe Covid-19. N Engl J Med 2020.

20. Joe W, Kumar A, Rajpal S, Mishra US, Subramanian SV. Equal risk, unequal burden? Gender differentials in COVID-19 mortality in India. J Glob Health Sci 2020;2.

